# Machine Learning Approaches to Identify Communities with High HIV Prevalence in Resource-Limited Settings using Social, Economic and Behavioral Data

**DOI:** 10.1101/2025.11.10.25339949

**Authors:** Masabho P. Milali, Frey B. Assefa, Sulani Nyimbili, Duncan K. Gathungu, Samuel Mwalili, Suilanji Sivile, Lloyd Mulenga, R. Scott Braithwaithe, Diego F Cuadros, Anna Bershteyn

## Abstract

**Background:** Identifying communities with high HIV prevalence is crucial for public health officials, researchers, and policymakers to effectively monitor the epidemic and evaluate interventions. Population-based HIV biomarker surveys face logistical challenges such as cost, need for personnel trained in specimen collection, specimen transport and processing, and participant reluctance to test due to factors such as stigma, history of recent testing, and the perception of being at low risk for HIV infection. This study explores the potential of identifying communities with high HIV prevalence using socio-economic, behavioral, and other community-level data in the absence of direct HIV biomarkers.

**Method:** Using the methods of Partial Least Squares (PLS) and Random Forests (RF), we developed machine learning models to predict HIV prevalence based on socio-economic and behavioral variables from Population-based HIV Impact Assessments (PHIA) surveys. Community HIV prevalence, derived from the PHIA biomarkers dataset, served as the dependent variable. Initially, models were trained to classify communities into <10% or ≥10% HIV prevalence categories. This procedure was repeated for prevalence thresholds of 5%, 7%, 15%, and 20%.

**Results:** PLS and RF achieved 79% and 80.5% accuracy, respectively, in classifying communities as having higher or lower HIV prevalence at a 10% threshold. At the 5%, 7%, and 15% thresholds, the models achieved similar accuracies, demonstrating consistent performance across varying thresholds, with RF slightly outperforming PLS. In both models, the variables that contributed most to classification included having a first intercourse experience before age 15, being uncircumcised, having a history of not using condoms, being in the lowest wealth quintile, experiencing physical or sexual violence, and having extramarital partners.

**Conclusions:** The study demonstrates that socioeconomic and behavioral variables can effectively predict community-level HIV prevalence using machine learning models. These insights have the potential to guide the distribution of HIV resources, particularly where direct community testing is infeasible, and to enhance understanding of the HIV epidemics.

## Introduction

The human immunodeficiency virus (HIV) remains a significant global health concern, particularly in those resource-limited regions where HIV prevalence is notably high (1,2). In response, health authorities have committed to UNAIDS’ 95-95-95 goals ensuring that 95% of people living with HIV know their status, 95% of those diagnosed receive antiretroviral therapy (ART), and 95% of those on ART achieve viral suppression (1,2).

While substantial progress has been made in improving the second and third 95-95-95 targets through improved accessibility of HIV testing and treatment, the first 95-95-95 target remains elusive in many settings because policymakers and implementers have inadequate information to identify areas where HIV testing interventions should be prioritized given limited resources (3). Identifying high-prevalence HIV communities is difficult for multiple reasons. First, logistical hurdles in collecting HIV biomarker data through traditional methods, such as blood tests, which are hindered by a scarcity of trained personnel and high costs in resource-limited settings (3–7). Second, current HIV surveys face limitations in reaching every community, typically sampling only 0.5–2% of communities in a PHIA survey (8). Third, personal factors such as stigma, recent testing, and perceptions of low risk for HIV infection contribute to reluctance in testing (5–7).

Accordingly, new HIV surveillance methods that do not requiring extensive resources are needed to address current limitations in identifying high-prevalence communities (3). While these new methods may not completely replace conventional approaches, complementary methods that utilize socio-economic and behavioral data can help address gaps by providing community-level estimates of HIV prevalence to guide interventions and allocate resources more effectively. In our data-rich world, fueled by digital connectivity and widely available socio-economic surveys like the Demographic and Health Surveys (DHS) (9), the Multiple Indicator Cluster Survey (MICS) (10), the Population-based HIV Impact Assessment (PHIA) surveys, along with censuses conducted globally, there is a wealth of information that can be leveraged. Remote surveys, such as questionnaires conducted by telephones, video calls or mobile platforms, can provide critical behavioral and socio-economic data at a lower cost and with broader reach. Combined with machine learning and artificial intelligence, these data sources offer a promising approach to estimate HIV prevalence across diverse communities without relying solely on conventional biomarker data.

In this study, we assessed the predictive ability of a novel approach for identifying communities with high HIV prevalence that leverages socio-economic and behavioral data from PHIA surveys along with machine learning techniques. We hypothesized that socio-economic and behavioral factors could serve as effective proxies for HIV prevalence in the absence of direct HIV biomarker data. To test this hypothesis, we trained models using random forest and partial least squares architectures, utilizing socio-economic, behavioral, and other community-level data.

## Methods

### Data

We conducted a retrospective study using de-identified data from the Population-based HIV Impact Assessment (PHIA) surveys conducted in Zambia and Kenya. No new data were collected, and the authors had no direct interaction with study participants. Access to the restricted PHIA datasets was granted through a formal data-use agreement in March 2024. All data were anonymized and aggregated at the community level prior to release, ensuring that no personally identifiable information was available to the authors.

The PHIA surveys are representative household and individual surveys that provide comprehensive data on socio-economic and behavioral factors related to HIV (8,11,12). Individual-level data are collected through structured interviews, capturing information on demographics, HIV risk factors, and household characteristics (8,11,12). Following the interviews, individuals who consent have a blood sample taken for detailed analysis of biological markers related to HIV infection, such as HIV serostatus, CD4 cell counts (which measure immune function), and viral load measurements (which assess the amount of HIV viruses in the blood), thereby creating a dataset commonly referred to as HIV biomarker dataset (8,11,12).

For the purposes of training our models, we generated three distinct datasets. The first dataset, comprising predictor variables from the Zambia PHIA surveys, will be referred to as ZAMPHIA for the remainder of the study. The second dataset includes variables from the Kenya PHIA surveys and will be referred to as KENPHIA. The third dataset, ZAKEPHIA, was created by combining common predictor variables from both the Kenyan and Zambian surveys, allowing us to train and test the model on a harmonized dataset that reflects shared predictors between the two countries.

### Generating Predictor Variables of Enumeration Areas

Using individual-level PHIA survey data, we derived predictor variables for each enumeration area (EA) by determining the proportion of individuals with specific characteristics within each EA, using the total number of eligible individuals as the denominator. We chose the total number of eligible individuals within each EA as the denominator to ensure that the derived proportions accurately reflect the prevalence of these characteristics within the entire population of interest in that specific area. For example, we calculated the proportion of circumcised individuals in an EA by dividing the number of circumcised males by the total male population within that EA, as this approach provides a direct measure of circumcision prevalence relative to the population that could have this characteristic (i.e., males), ensuring that the predictor variables are representative and comparable across different EAs. In addition, we included the distance from each EA to the nearest health facilities and major transit roads. The complete list of all generated predictor variables is provided in Table S1 of the appendix. An EA is a geographic area with a radius of 200 meters in urban settings and 1,000 meters in rural settings, serving as the primary sampling unit to systematically survey the target population efficiently and to gather sufficient data (13).

### Computing HIV Prevalence of Enumeration Areas

HIV prevalence at each EA level was computed using the PHIA adult biomarkers dataset (8). This was done by dividing the number of people who tested positive for HIV by the total number of individuals who consented to have their blood sample taken for biomarker analysis in that EA. Individuals in the EA who did not have an HIV status recorded were not included in the computation of the HIV prevalence for that EA.

### Distance of each EA to health facilities

We calculated the distances from each EA to the nearest health facilities using geo-coordinates. The geo-coordinates of each EA were provided by the PHIA surveys, while those for health facilities in Kenya and Zambia were obtained from a published manuscript. This additional data enabled us to assess the impact of access to healthcare on community-level HIV prevalence.

### Model Training to estimate HIV prevalence class of an EA

Initially, EAs within each dataset (ZAMPHIA, KENPHIA, and ZAKEPHIA) were categorized into two groups based on HIV prevalence: those with less than 10% were labeled ’coldspots’ (0), and those with 10% or higher were labeled ’hotspots’ (1). After categorization, we merged and then randomized the EAs within each dataset into training (70% of the data) and testing (30% of the data) sets, where the number of EAs (N) is 511 for ZAMPHIA, 798 for KENPHIA, and 1,309 for ZAKEPHIA (Figure 1). This randomization ensures that the models are trained and tested on distinct subsets of data, reducing the risk of overfitting.

**Figure 1.**
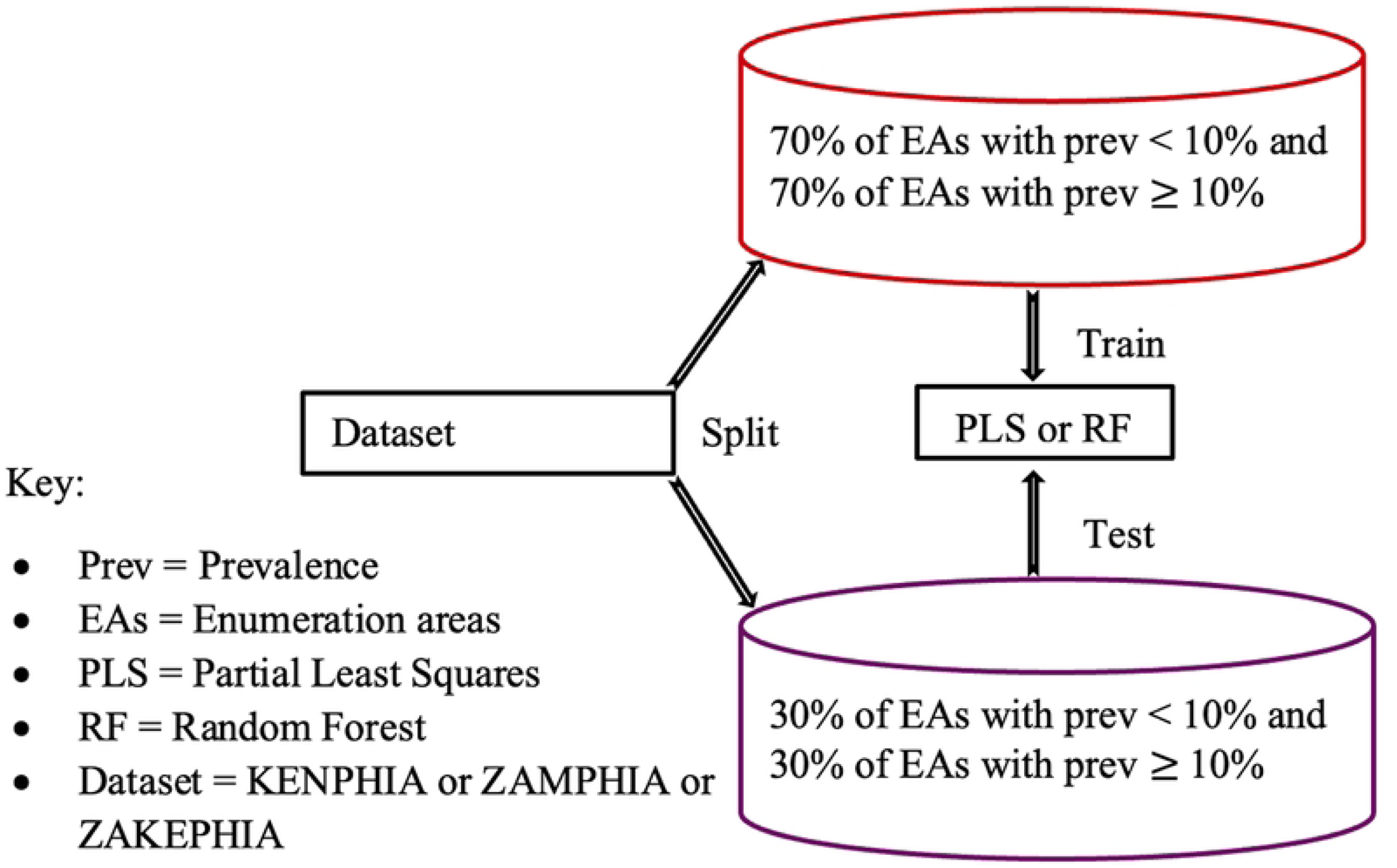
Illustration of how the dataset was divided into training and test sets.

Each dataset was used to train a 15-component Partial Least Squares (PLS) model with ten-fold cross-validation, and a Random Forest with 100 trees and a maximum depth of 4 (RF). The selection of 15 components for the PLS model was based on the percentage of variance explained in the response variable as a function of the number of components, a standard technique used to assess PLS performance in feature reduction (Figure 2). This approach ensures the preservation of valuable information in the dataset while maintaining model accuracy and avoiding overfitting (14–16).

**Figure 2.**
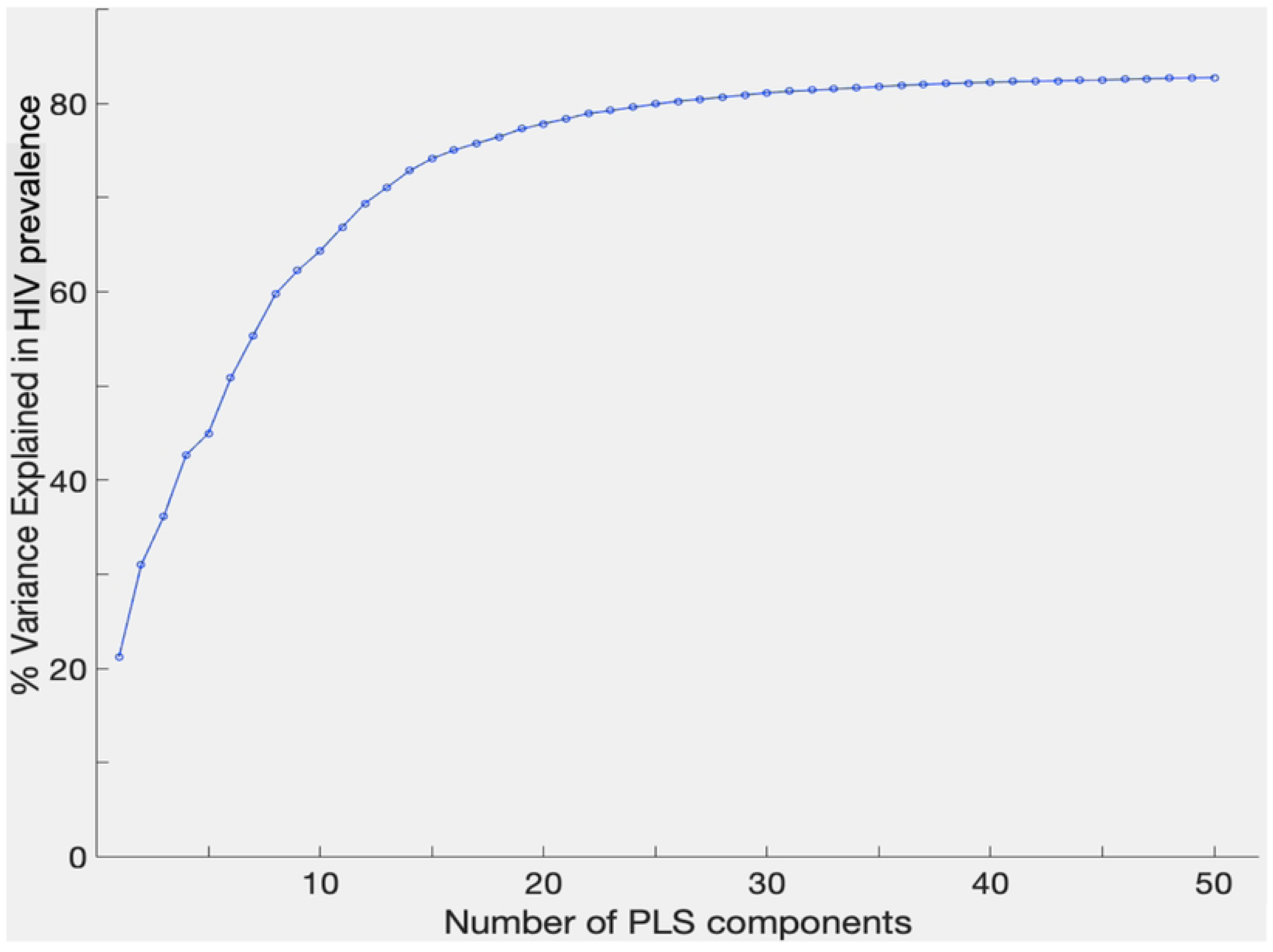
Percentage of variance in community HIV prevalence explained by the number of Partial Least Squares (PLS) components. This figure informs selection of an optimal number of components to explain variance in HIV prevalence.

The decision to train trees with a maximum depth of 4 in the RF104 model was guided by the cross-validation method, a common approach for selecting the optimal depth by training trees of various depths, evaluating their performance on validation data, and determining which depth minimizes error on unseen data (Figure 3) (17).

**Figure 3.**
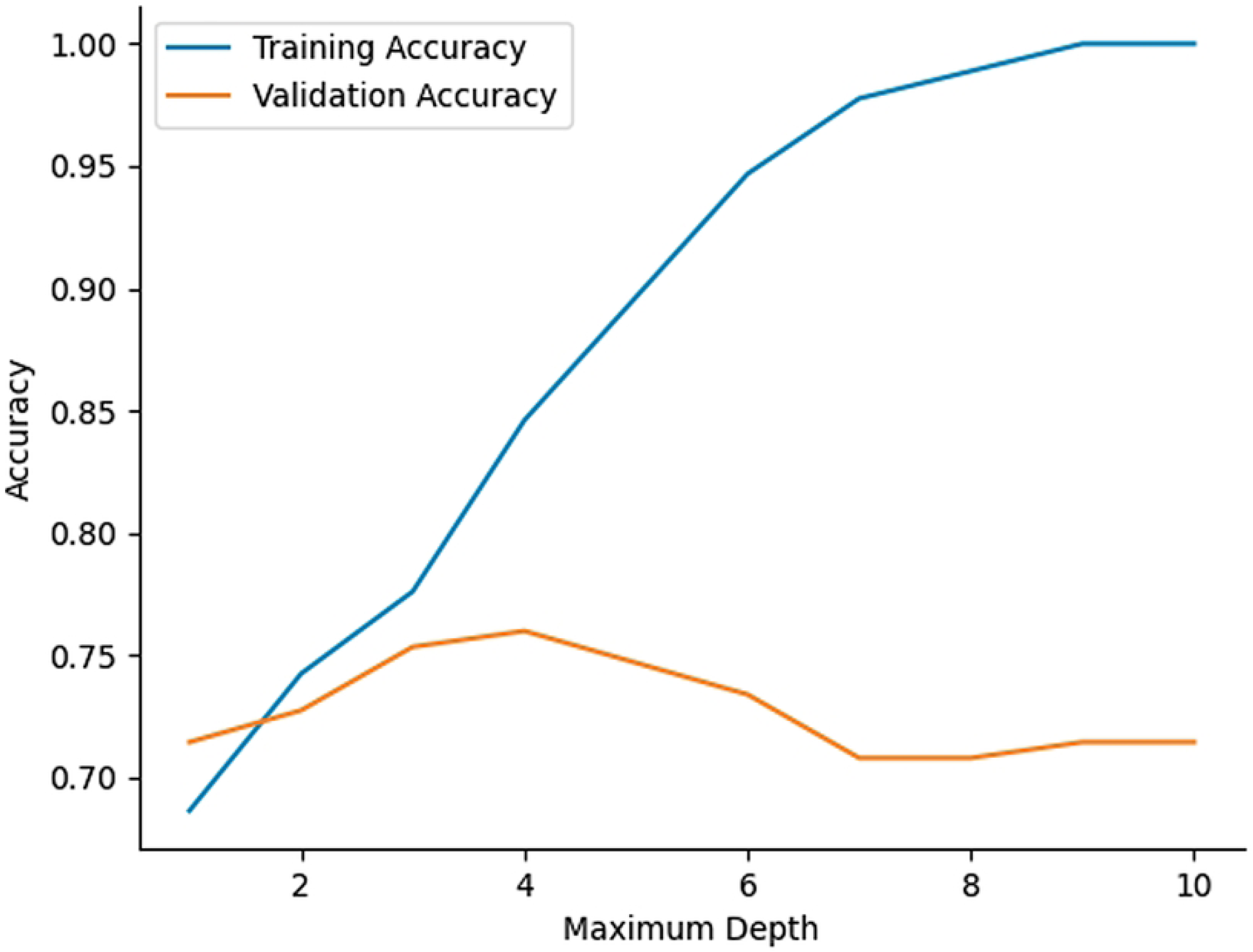
Cross-validation approach for selecting the optimal tree depth. This figure shows training and validation accuracy across various tree depths. The training accuracy increases with depth, indicating the model’s capacity to fit the training data better at higher depths. However, validation accuracy plateaus and begins to decline slightly, suggesting overfitting at greater depths. The optimal depth is chosen to maximize validation accuracy while minimizing error on unseen data.

For model interpretation, EAs were classified based on the model’s probability score, a continuous variable ranging from 0 to 1. EAs with a probability score below the 0.5 threshold were classified as having less than 10% HIV prevalence, while those with a score of 0.5 or above were classified as having 10% or higher. This probability score is derived from the model’s internal calculations, using input data, and represents the likelihood that a given EA meets the criteria for higher or lower HIV prevalence. Specificity, sensitivity, precision, and accuracy for both PLS and RF were calculated using Equations 1, 2, 3, and 4. Sensitivity of the model is the ability to classify EAs correctly, which have HIV prevalence greater than or equal to 10% (assumed to be HIV hotspot), and specificity is the ability of the model to classify EAs correctly which are less than 10% HIV prevalence (assumed to be HIV coldspot) (18–20).

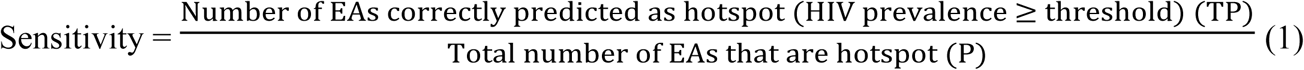

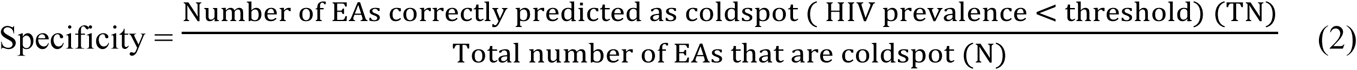

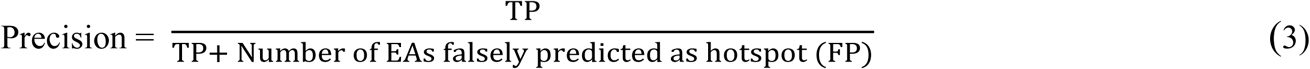

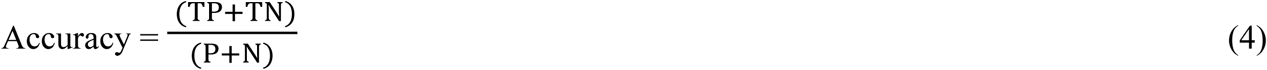

The process of randomly splitting the data into training and test sets, followed by training and testing the models, was repeated ten times - a method known as Monte Carlo cross-validation (21–23). To test the robustness of the model accuracy on the HIV prevalence threshold, we repeated the model training and testing similar to the approach with the 10% threshold using additional thresholds of 5%, 7%, and 15%.

To evaluation the extrapolation capability of the PLS and RF models on datasets with different sample characteristics from the training sets, we conducted cross-testing by applying the model trained on ZAMPHIA to KENPHIA and vice versa. Four testing scenarios were implemented. In Test A, the model was trained on the ZAMPHIA dataset and tested on the KENPHIA dataset, while in Test B, the model was trained on the KENPHIA dataset and tested on the ZAMPHIA dataset. Test C involved training the model on both the ZAMPHIA and KENPHIA datasets and testing it on the ZAMPHIA dataset, whereas in Test D, the model was trained on both datasets and tested on the KENPHIA dataset.

### Analysis of Variable Importance

After model training and confirming acceptable accuracy, we analyzed the importance of each community variable in predicting HIV prevalence. In the PLS model, we applied the Variable Importance in Projection (VIP) score to identify features that significantly influenced model accuracy. This score, frequently utilized in analyses of large datasets characterized by multicollinearity, is determined by a weighted sum of the squares of the variable weights, which reflects their importance within each component (14,24). Variables with a VIP score above 1 are regarded as more important, whereas those with scores below 1 are considered less important (14,24).

In the Random Forest model, variable importance was evaluated using the Gini impurity index (25,26). Gini impurity is defined as 1 - ∑^𝑘^_*i*=1_ 𝑝^2^_*i*_ , where 𝑝_𝑖_ represents the proportion of observations belonging to class *i* within the dataset (26). It spans from 0 to 1, with 0 indicating complete purity – where all observations belong to a single class, and 1 denoting maximum impurity, with observations evenly distributed across various classes(25–28). This index enables the decision tree algorithm to select the variable that, when used to binary split observations in the dataset, ensures a substantial decrease in Gini impurity, hence creating the most homogeneous branches. Each variable’s importance was determined by its contribution to the reduction of Gini impurity across all splits it generates within the tree. The reduction in Gini impurity for each split was calculated and aggregated for each variable (feature) throughout the tree’s construction. After the decision tree for hotspot classification was built, the aggregate reduction for each variable was normalized against the sum of reductions across all variables to obtain a standardized measure of importance (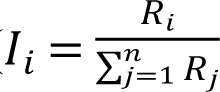, where 𝐼_𝑖_ is normalized importance of feature *i*, 𝑅_𝑖_ is aggregate reduction for feature *i*, and 𝑅_𝑗_ is the aggregate reduction for the j^th^ feature within a decision tree) (29). To establish the final importance score for each variable in a random forest model, we computed the average of these normalized scores across 100 independently trained trees.

The variable with the highest average reduction in Gini impurity is identified as the most important feature in the model, as its role as a splitting variable resulted in the greatest improvement in node purity. In another words, using this variable to divide the data created child nodes that were more uniform in class distribution compared to the parent node.

## Results

Overall, all both PLS and RF models demonstrated robust accuracy (76 ± 5%) across all datasets, with notable variations at different thresholds. When model performance was compared, RF models consistently outperformed PLS models in terms of accuracy and sensitivity across all datasets, particularly at a 15% HIV prevalence threshold, where RF models achieved the highest accuracy and sensitivity. Specificity was generally higher in RF models, indicating an ability to correctly identify communities with lower HIV prevalence. Precision was variable, with PLS models sometimes outperforming or matching RF models, especially at lower prevalence thresholds (Figure 4).

**Fig 4.**
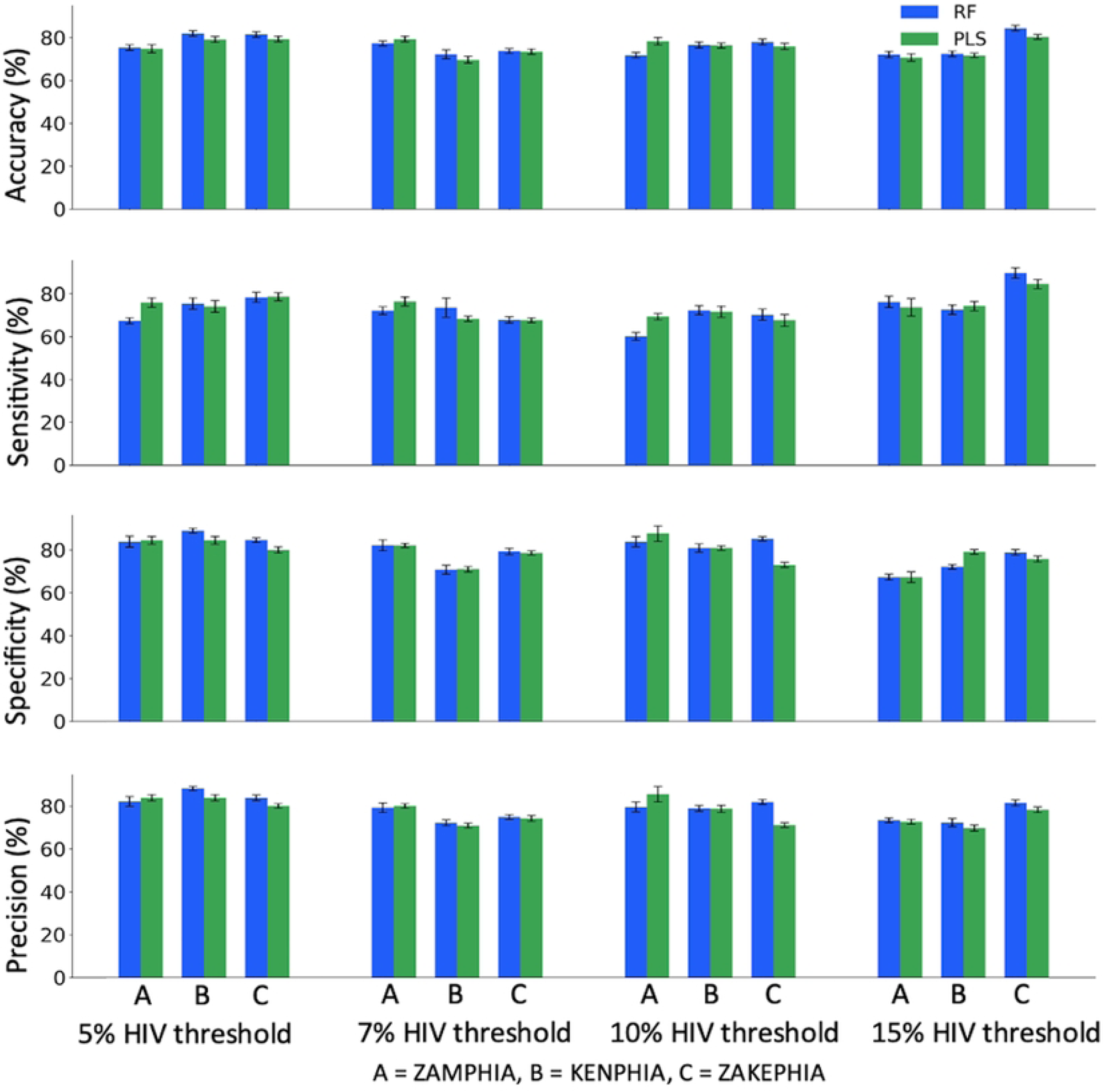
Performance of Random Forest and Partial Least Square models across different datasets and HIV prevalence thresholds. Models were trained and validated using three distinct datasets: Zambia PHIA surveys (A), Kenya PHIA surveys (B), and Zambia-Kenya PHIA surveys (C). Performance metrics (accuracy, sensitivity, specificity, and precision) are displayed for HIV prevalence thresholds set at 5%, 7%, 10%, and 15%. Each bar represents the average metric score across ten Monte Carlo cross-validation iterations, illustrating the models’ consistency and reliability in identifying HIV prevalence class in the study regions.

Furthermore, although RF models generally achieved higher AUCs than PLS models, both RF and PLS models exhibited consistently high area under the curve (AUC) values, indicating stable performance in distinguishing communities with high and low HIV prevalence across various probability thresholds – a recommended performance test for binary classifiers (Table 1 and Figures S1 and S2 in the appendix).

**Table 1.** Area under the curve (AUC) values for Random Forest (RF104) and Partial Least Squares (PLS) models trained on Zambia (ZAMPHIA) and Kenya (KENPHIA) PHIA surveys at HIV prevalence thresholds of 5%, 7%, 10%, and 15%, with model performance at each threshold evaluated across various probability scores.

| Dataset | Model type | HIV prevalence threshold |  |  |  |
| --- | --- | --- | --- | --- | --- |
|  |  | 5% | 7% | 10% | 15% |
| ZAMPHIA | RF104 | 0.88 | 0.86 | 0.85 | 0.78 |
|  | PLS | 0.86 | 0.81 | 0.86 | 0.78 |
| KENPHIA | RF104 | 0.79 | 0.80 | 0.91 | 0.97 |
|  | PLS | 0.75 | 0.79 | 0.85 | 0.93 |

In cross-testing models on datasets from different geographic regions, the models achieved an overall accuracy of approximately 65% across all tests and thresholds (Figure 5). Test A, trained on ZAMPHIA and tested on KENPHIA, and Test B, trained on KENPHIA and tested on ZAMPHIA, exhibited moderate accuracy but generally lower sensitivity and specificity. Tests C and D, which were trained on both the ZAMPHIA and KENPHIA datasets and tested on ZAMPHIA and KENPHIA, respectively, demonstrated improved performance. In addition, RF models slightly outperformed PLS models in most cases, particularly in sensitivity and specificity.

**Fig 5.**
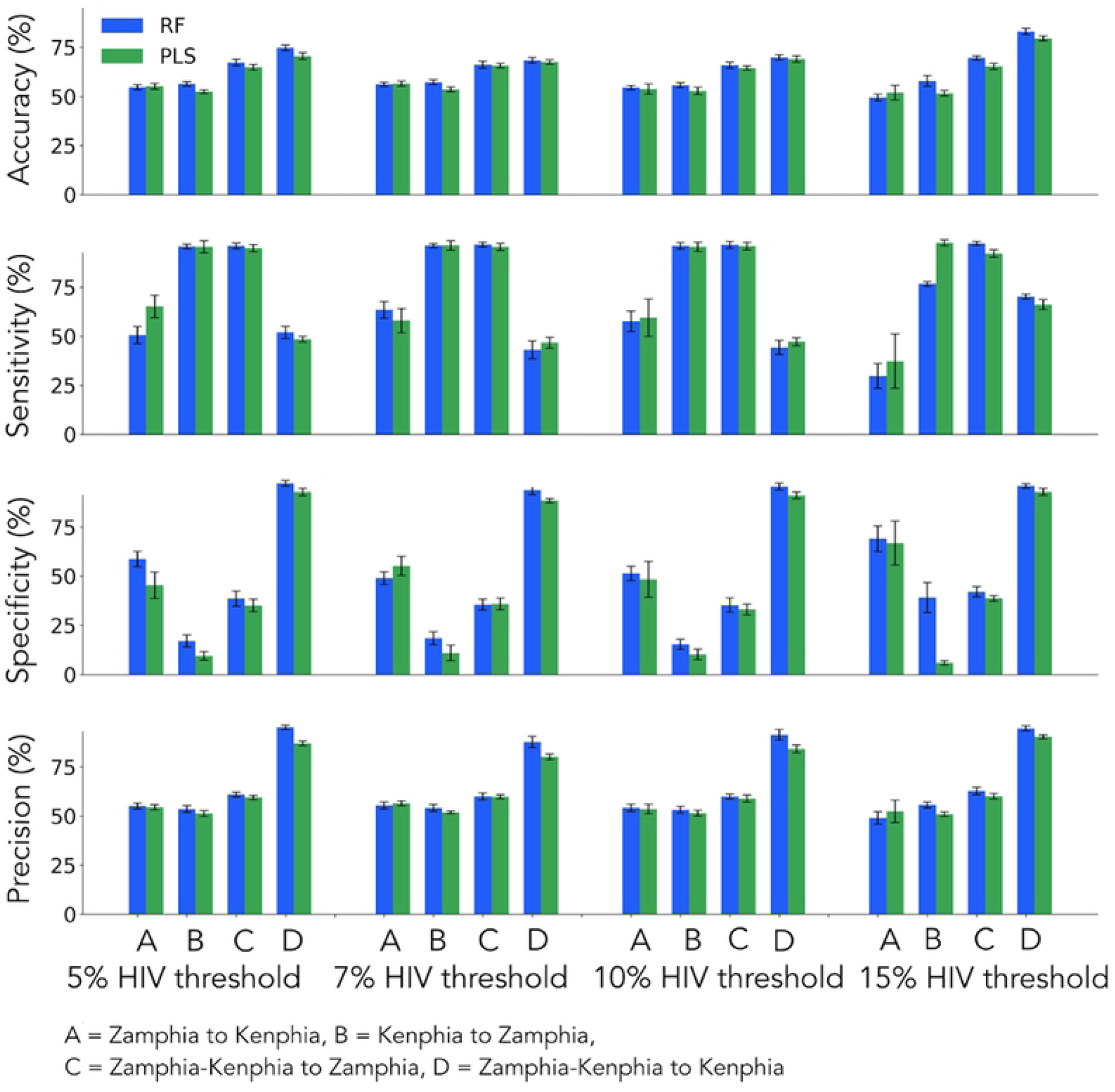
Performance of Random Forest and Partial Least Squares models on cross-testing across three datasets: A) Trained on ZAMPHIA and tested on KENPHIA, B) Trained on KENPHIA and tested on ZAMPHIA, C) Trained on both ZAMPHIA and KENPHIA and tested on ZAMPHIA, D) Trained on both ZAMPHIA and KENPHIA and tested on KENPHIA. This test allows for a direct comparison of how well models trained on one country’s data predict HIV prevalence in the other, as well as how combined datasets influence prediction accuracy.

The analysis identified important variables that consistently influenced the performance of both RF and PLS models across different thresholds. In the KENPHIA dataset, important variables included the proportion of individuals entering relationships for financial support, the proportion of uncircumcised males, and individuals who had experienced physical or sexual violence. For the ZAMPHIA dataset, important variables included the proportion of women married before age 18, individuals in the lowest wealth quintile, and the proportion of individuals with secondary-level education. Behavioral variables, such as the proportion of individuals not using condoms during sexual encounters with short-term partners, were also important in accurately classifying community HIV prevalence. A comprehensive list of variables and their categories can be found in (Figure 6).

**Fig 6.**
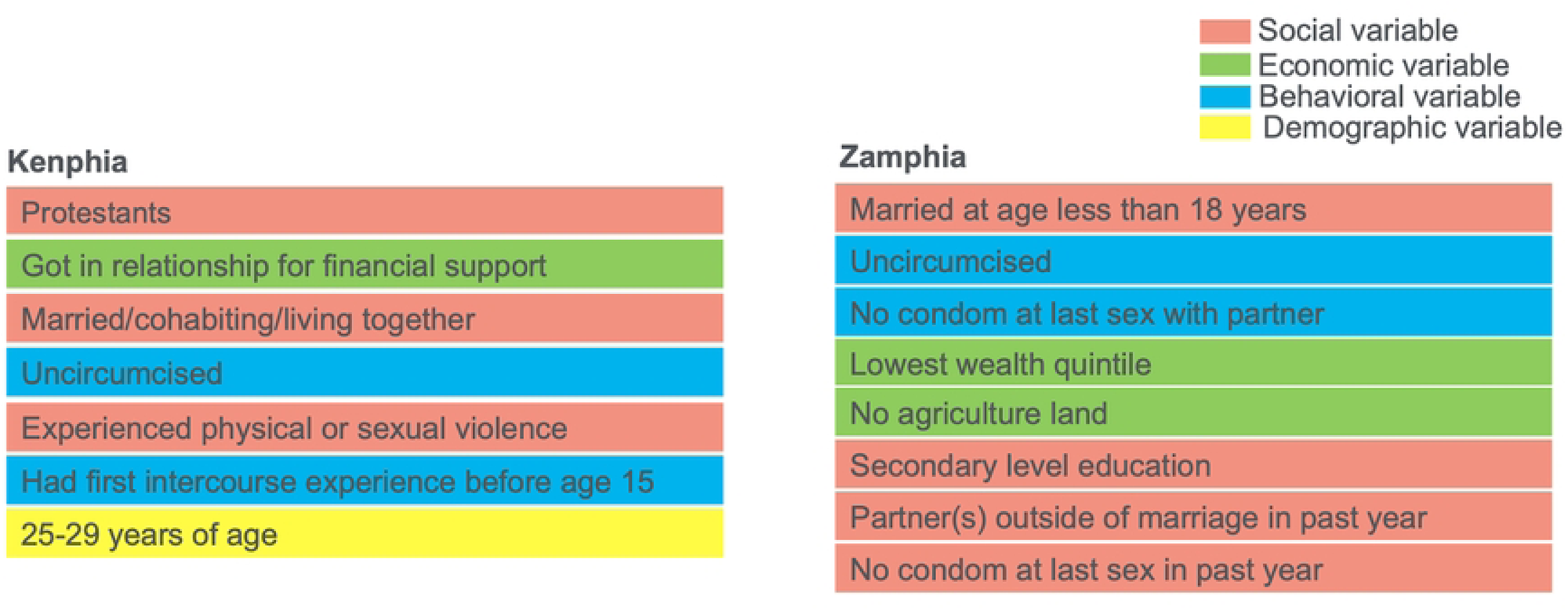
Important variables that influenced models’ performance across 5%, 7%, 10% and 15% HIV prevalence thresholds. Variables are categorized into social (orange), economic (green), behavioral (blue), and demographic (yellow) groups. Each category highlights factors like marital status, circumcision status, sexual behavior, and age, which are critical in analyzing HIV prevalence across the surveyed populations.

## Discussion

This study developed and tested two models using socio-economic and behavioral data from the Zambia and Kenya PHIA surveys to identify communities with higher HIV prevalence. RF and PLS models achieved accuracies between 79% and 81% for classifying high- vs. low-prevalence communities. Training and validating models across multiple HIV prevalence thresholds (5%, 7%, 10%, and 15%) provided a nuanced understanding of the key variables influencing their accuracy in estimating community HIV prevalence. Variables such as marital status, sexual behavior, and financial status – including early sexual debut, lack of circumcision, inconsistent condom use, low income, experiences of violence, and extramarital relationships consistently emerged as important predictors across all thresholds.

Our findings demonstrate the potential of machine learning to approximate HIV prevalence using non-biological data. The use of Monte Carlo cross-validation methods (21,22) in this study demonstrated the stability and reliability of our models across diverse training and testing scenarios, suggesting that the findings are not artifacts of specific samples or model specifications. As recommended for binary classifiers, ROC curves were generated to provide a detailed analysis of the models’ performance across different probability thresholds, revealing their capacity to distinguish between varying levels of HIV prevalence (30). Our study further demonstrated that RF models generally provided higher accuracy and sensitivity compared to PLS models. This might be attributed to the RF model’s ability to handle complex interactions and non-linear relationships among predictors more effectively than PLS models (31,32). The high sensitivity of RF models is particularly advantageous in public health contexts, as it ensures that high-risk areas are less likely to be missed by the surveillance programs. Meanwhile, the robust specificity exhibited by the RF models helps minimize false alarms - areas falsely identified as high-risk - which can lead to more targeted and cost-effective interventions.

The cross-testing results reveal that models trained on datasets from a single geographical setting, such as ZAMPHIA or KENPHIA, exhibited moderate accuracy (approximately 65%) but lower sensitivity and specificity when applied to data from a different region (Figure 5). This outcome highlights the inherent limitations of a single-region training, as such models are tailored to the specific socio-economic and behavioral characteristics of the region and may not fully capture the diversity of factors influencing HIV prevalence elsewhere. In contrast, models trained on combined datasets from both ZAMPHIA and KENPHIA demonstrated consistently improved performance across accuracy, sensitivity, specificity, and precision metrics. These findings underscore the importance of integrating data from multiple regions to enhance generalizability and robustness, particularly for applications in geographically diverse settings. While these findings demonstrate the potential of cross-regional data integration, further refinement is needed to ensure robust performance across diverse contexts before considering large-scale applications. For instance, incorporating additional variables reflecting contextual factors or leveraging advanced modeling techniques such as ensemble learning or deep learning could enhance model accuracy and generalizability.

Our study is not the first to employ machine learning algorithms in the HIV domain to demonstrate the potential of models trained on socio-economic and behavioral data for HIV screening. Previous studies (33–35) have shown that gradient boosting trees (36) and the XGBoost algorithm (37), when applied to socio-behavioral features, can effectively identify individuals at increased risk of infection in East and Southern African countries, supporting the findings of our study. While identifying individuals at increased risk of acquiring HIV, as reported by previous studies (33–35), is useful and important, identifying communities with higher HIV prevalence, as demonstrated in this study, provides a comprehensive view of the epidemic at the population level. This approach reduces stigma, informs resource allocation, and enables broader, more effective public health interventions. It complements individual-level data by creating a more complete picture of the HIV epidemic, which is necessary for tackling root causes and preventing future transmission on a larger scale. The variables most predictive of HIV prevalence in our study are consistent with existing literature linking these variables to increased HIV risk (38–43). These variables may have applications in developing targeted public health interventions, addressing the underlying socio-economic and behavioral drivers of HIV transmission.

Despite the promising results of this study, it has several important limitations. First, a key constraint is the infrequent availability of updated PHIA survey data, which are typically conducted approximately every five years and often experience additional delays before dataset release. Consequently, the analyses may not always reflect the most current HIV epidemiology. The key variables driving model accuracy in estimating community prevalence may change over time due to evolving HIV programs and interventions. Therefore, retraining the models with more recent PHIA data, when available, will be necessary to ensure they incorporate the latest variables influencing HIV transmission and maintain performance. Second, although PHIA surveys are conducted in countries beyond Kenya and Zambia, our study did not include data from these additional regions. Incorporating PHIA surveys from other countries could provide additional insights of the factors influencing HIV prevalence in sub-Saharan regions, and is an important area for future work. Third, our study did not utilize data from other extensive surveys such as the Demographic and Health Surveys (DHS) and national censuses, which also contain valuable socio-economic and behavioral data. This limitation reduced the breadth of data available to train our model more comprehensively. Like other machine learning models, our approach would benefit from more data points per community (EA), which would allow for the use of regression models instead of classification models. With a larger amount of data per EA, regression models could be used to estimate a continuous range of HIV prevalence values, rather than simply classifying communities as having high or low prevalence. This approach would allow for more nuanced insights into prevalence levels across communities, but it requires a sufficient number of data points for accurate training.

Further research could focus on addressing our limitations above and explore integrating additional data layers, such as mobility data, which could provide deeper insights into the dynamics of HIV transmission. Additionally, extending this modeling approach to other infectious diseases could enhance the generalizability and utility of our findings across different public health challenges.

## Conclusion

This study demonstrates the potential of machine learning models trained on socio-economic and behavioral data to enhance HIV surveillance by enabling remote, data-driven estimation of community HIV prevalence. This approach allows for real-time monitoring of emerging HIV hotspots and supports rapid intervention in areas where conventional biomarker-based methods are financially and/o logistically challenging, particularly in resource-limited settings. By leveraging readily available survey data, this model provides a scalable and low cost complement to traditional surveillance strategies.

## Ethics approval and consent to participate

This study involved secondary analysis of de-identified data obtained from the Population-Based HIV Impact Assessment (PHIA) program. Ethical approval for the original PHIA surveys was granted by the respective national and institutional review boards, including the Zambia National Health Research Ethics Board (Ref: MH/101/23/10-1), the Tropical Diseases Research Centers Ethics Review Committee (Ref: STC/2015/9) for ZAMPHIA, the Kenya Medical Research Institute–Science and Ethics Research Unit (KEMRI-SERU-592) for KENPHIA, the U.S. Centers for Disease Control and Prevention (CDC-IRB-7094), and the Columbia University Institutional Review Board (IRB-AAAR7792 (Y05M01)). All original PHIA participants provided written informed consent prior to participation. For the present analysis, no new data were collected and no direct interaction with participants occurred; therefore, additional ethics approval was not required. Access to the restricted PHIA datasets was granted through a formal data-use agreement with the PHIA Data Repository (https://phia-data.icap.columbia.edu).

## Consent for publication

Not applicable

## Availability of data and materials

The data that support the findings of this study are available from the Population-Based HIV Impact Assessment (PHIA; https://phia-data.icap.columbia.edu/), but restrictions apply to the availability of these data, which were used under license for the current study and so are not publicly available. We sought and were granted permission to use the core data set for this analysis by PHIA.

## Authors’ contributions

MPM performed the analysis, prepared the figures, and wrote the first draft of the manuscript. FA preprocessed the data used for the analysis. All authors reviewed the manuscript.

## Data Availability

The data that support the findings of this study are available from the Population-Based HIV Impact Assessment (PHIA https://phia-data.icap.columbia.edu/), but restrictions apply to the availability of these data, which were used under license for the current study and so are not publicly available. We sought and were granted permission to use the core data set for this analysis by PHIA.

## Acknowledgments

Not applicable

## Supporting information

**Table S1.**
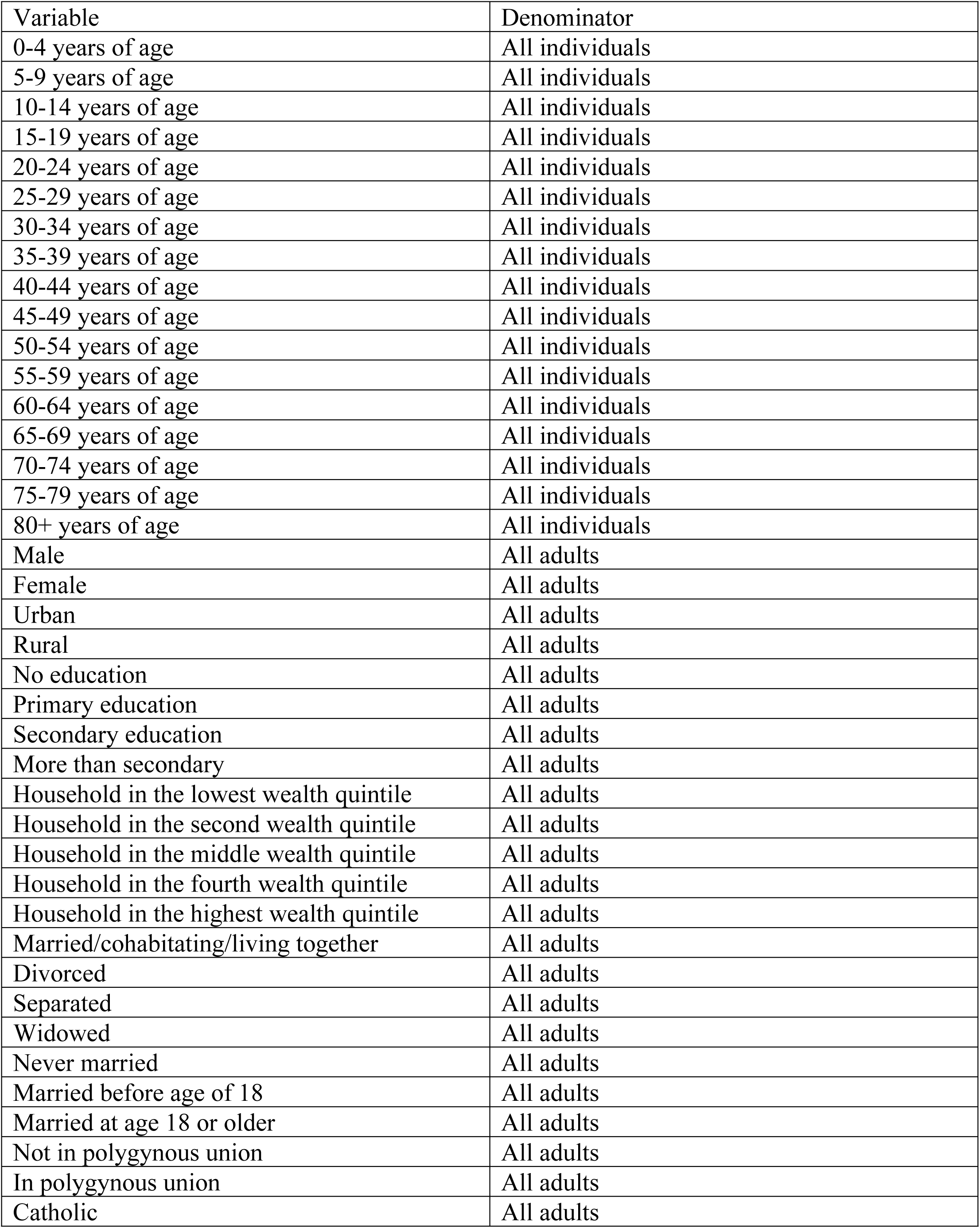
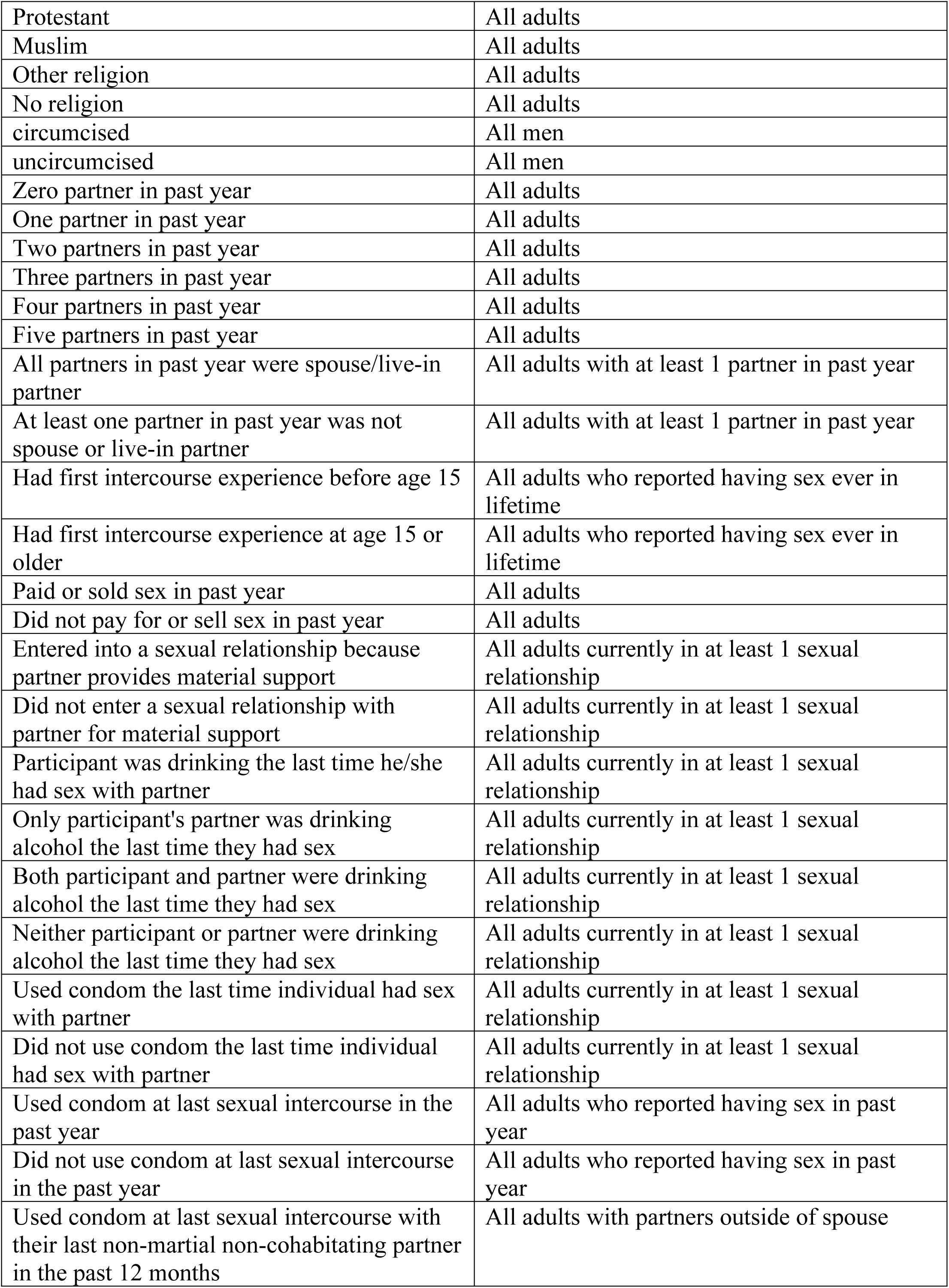
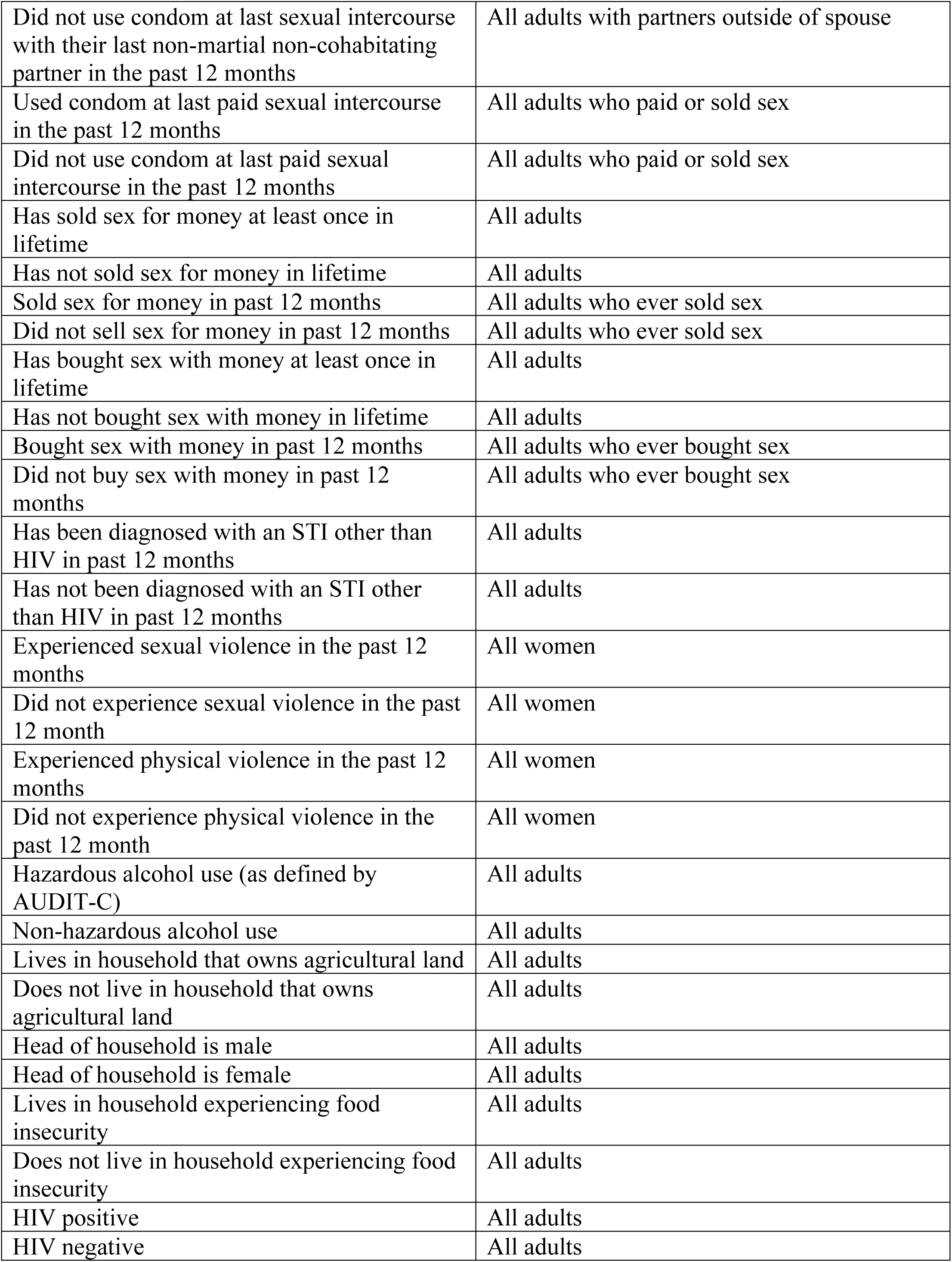
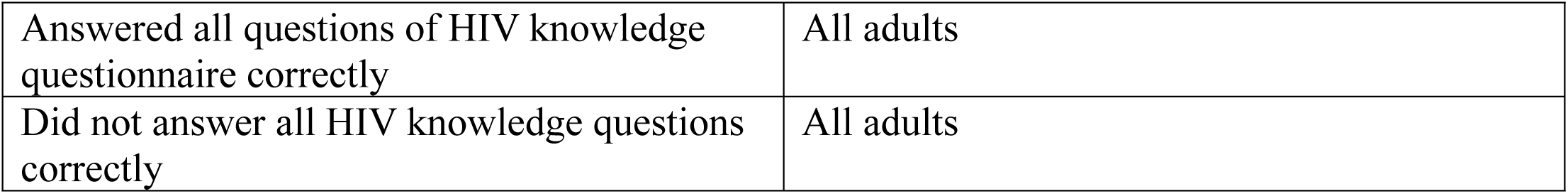
The complete list of all generated predictor variables from PHIA surveys used for the model training and testing process.

**S1 Figure.**
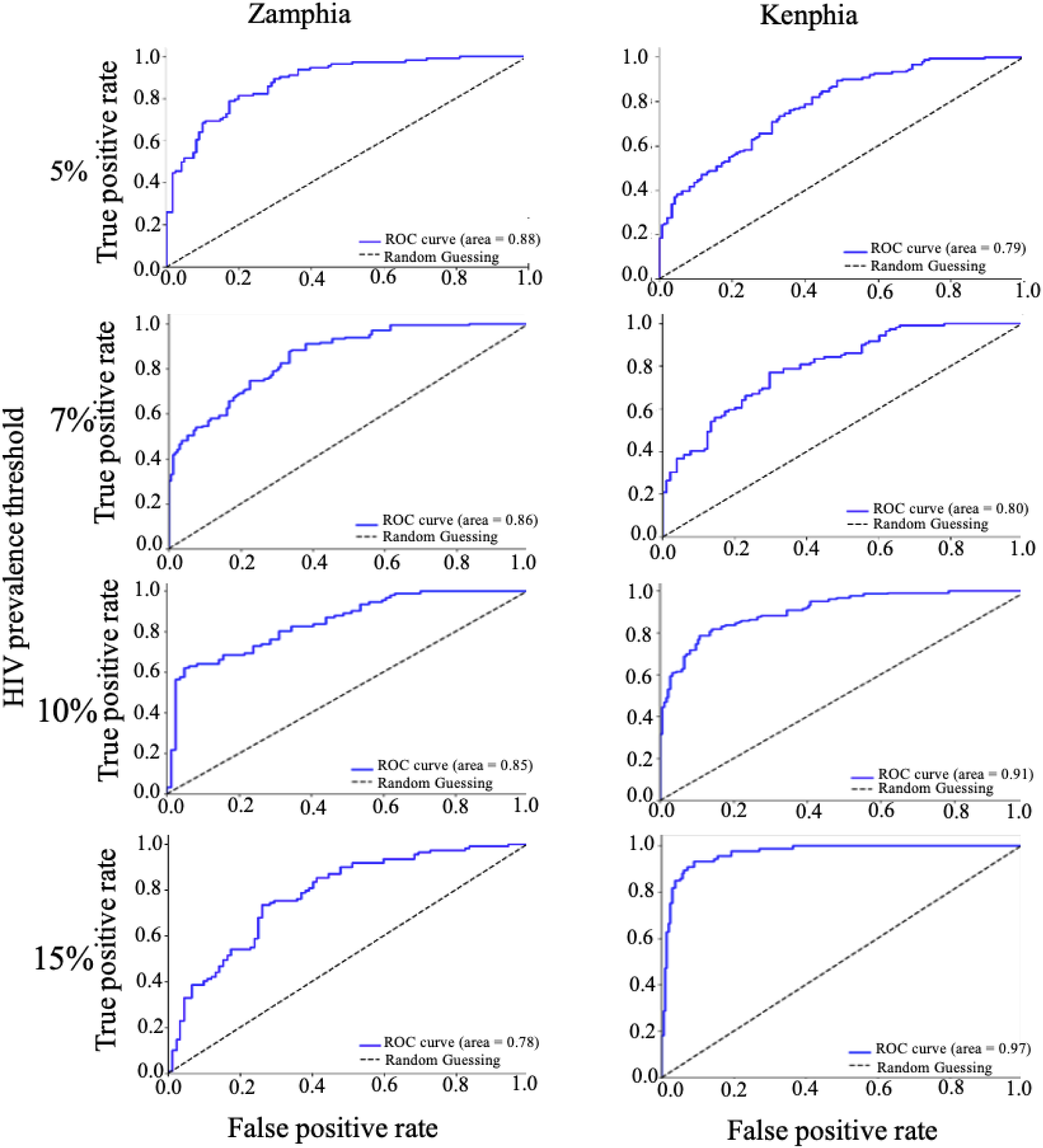
Receiver Operating Characteristic (ROC) curves for Random Forest models trained on Zambia and Kenya PHIA surveys at HIV prevalence thresholds of 5%, 7%, 10%, and 15%. The curves represent the trade-off between sensitivity (true positive rate) and specificity (false positive rate) across various threshold settings. A diagonal dashed line indicating random guessing (ROC curve area = 0.5) serves as a baseline for comparison. The area under the ROC curve (AUC) for each scenario is provided in the legend within each panel, illustrating the model’s ability to accurately classify the HIV prevalence based on the selected threshold.

**S2 Figure.**
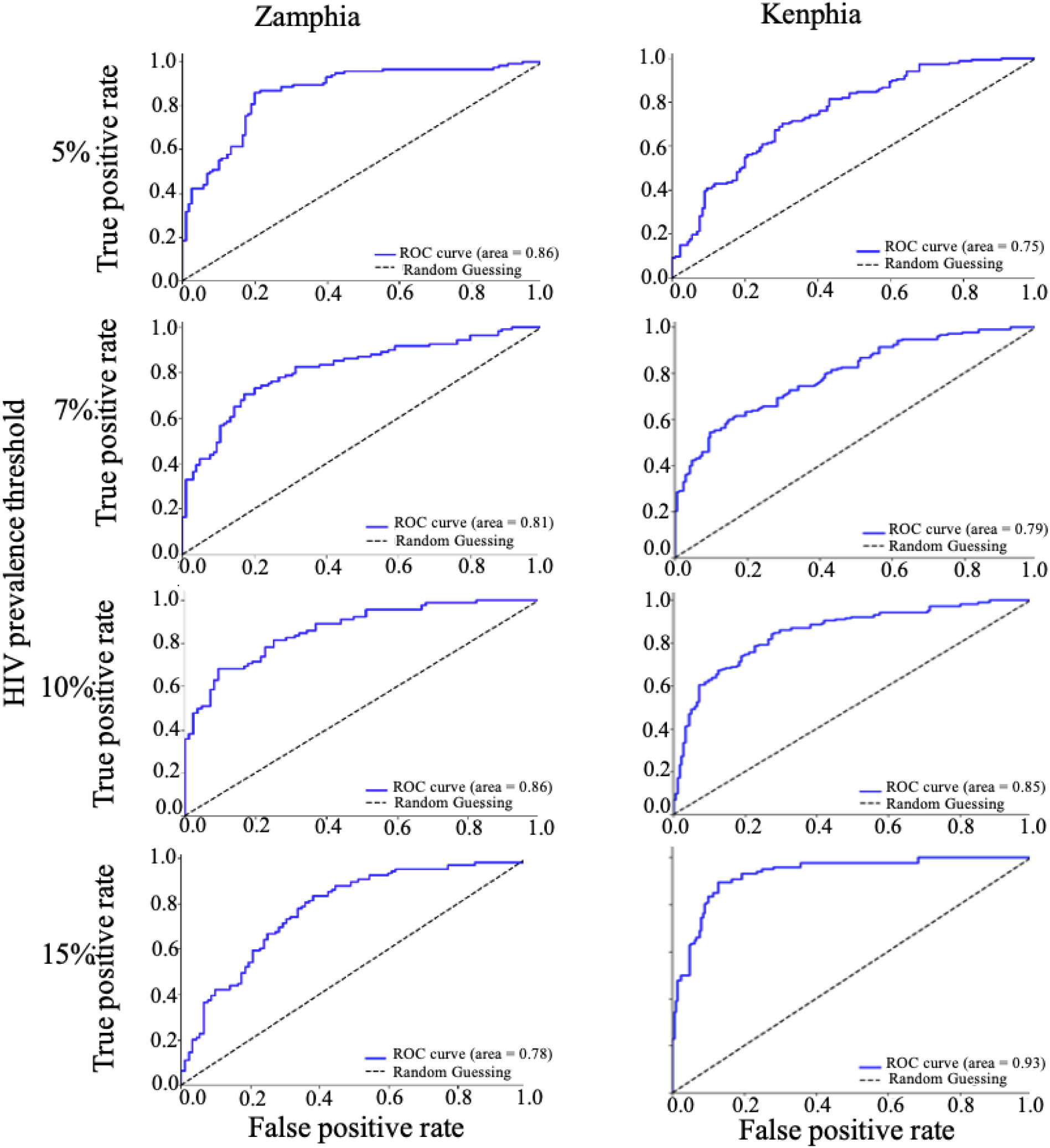
Receiver Operating Characteristic (ROC) curves for Partial Least Squares models trained on Zambia and Kenya PHIA surveys at HIV prevalence thresholds of 5%, 7%, 10%, and 15%. The curves represent the trade-off between sensitivity (true positive rate) and specificity (false positive rate) across various threshold settings. A diagonal dashed line indicating random guessing (ROC curve area = 0.5) serves as a baseline for comparison. The area under the ROC curve (AUC) for each scenario is provided in the legend within each panel, illustrating the model’s ability to accurately classify the HIV prevalence based on the selected threshold.

